# COMPETENCES OF LOWER COMMUNITY HEALTH CENTRE LEADERS IN ANNUAL HEALTH WORK PLANS AND ITS INFLUENCE ON DISTRICT PERFORMANCE IMPROVEMENT IN BUSOGA SUB-REGION: A RETROSPECTIVE STUDY

**DOI:** 10.1101/2024.12.07.24318647

**Authors:** Kharim Mwebaza Muluya, Gangu David Muwanguzi, Aremu Babatunde, Naziru Rashid, Irene Wananda, Jonah Fred Kayemba, Collin Ogara, Musa Waibi, John Francis Mugisha, Peter Waiswa

## Abstract

**Background:** Lower-level community health centres play a crucial role in the delivery of primary healthcare services, and the competences of their leaders can significantly influence district health performance.

**Objective:** The study assessed the influence of competences of lower community health centre leaders in annual health work planning on the district performance improvement. Staffs are recruited based on staffing standards and expected to participate in district health planning cycles, and use implementation manuals in developing annual health work plans to improve district health performance.

**Methods:** A retrospective (case control) study design was employed to understand health centre performance data across various districts in the Busoga sub-region. There was a comparison of performance between the worst performing (case) and best performing (control) districts in the region according to the Annual Health Sector Performance reports from 2017/18 financial year to 2021/2022. Data was collected between 17^th^ July, 2024 and 23^rd^ August, 2024. Statistical analysis of data from 12 health centres in the case and 12 in the control was conducted using STATA *version 16* to determine competences of lower community health centre leaders that influence district performance.

**Results:** The study found that the district performance in annual health work planning was not good in both the case and control groups (26.4% and 47.2% respectively). Only three competences variables were statistically significantly associated with improved district health performance. Health centres that (1) met staffing levels in accordance with public facility standards (χ^2^, 7.756; *p-value, 0.005\**), (2) staff attended district planning cycle meetings (χ^2^ 16.713; *p-value, 0.001\**), and (3) health centres utilized four or more implementation manuals for the development of their annual health work plans (χ^2^ 43.333; *p-value, 0.001\**) demonstrated statistically significant improvements in performance. These competences enabled more effective service delivery, better alignment with district health priorities, and the implementation of comprehensive, evidence-based interventions.

**Conclusion:** The competences of lower community health centre leaders is held together with health centres meeting staffing standards, their participation in district planning cycles, and the utilization of implementation manuals and are critical for improving district health performance in the Busoga sub-region. Strengthening these competences through targeted supportive meetings and capacity-building initiatives is recommended to enhance the overall effectiveness of health service delivery at the district level.

## Introduction

Annual Work Plans (AWP) means the annual work program to be prepared by the recipient during each calendar year, including a program of activities and their performance in the concluding year followed by a forecast of the schedule of activities proposed for inclusion in the following fiscal year (Ntirandekura *et al.,* 2022). It is a set of successive activities over a period of one year, interconnected and which contribute to the same broader aim and created to oversee planned activities, and expected results (Ntirandekura *et al.,* 2022). An annual work plan can be as complex or as simple as the organization wants it to be and the policy-making levels set the barometers (Ntirandekura *et al.,* 2022). Organizations worldwide, therefore, are in the fray of making annual work plans for the better implementation of their programs.

In the Uganda Health Sector, the Annual Health Work Plan is an annual recurring activity performed by health centre II, Health Centre III, and other upper-level government health facilities and these plans provide suitable performance improvement score guides and means of verification according to the Local Government Management of Service Delivery Performance Assessment Manual provided for the higher local governments (2020). Performance improvement is enticed by managing health centre performance gaps to provide good services to the needy population. Lower community health centres have been given the opportunities to entangle the autonomy as enablers of community members to enjoy better services as emphasized by the government (Lotunani *et al*., 2014). The lower community health centres include health centres II, III and IV in the lower local governments (LLGs). Granting broader power to the lower health centres needs good, harmonious coordination and management at different levels including national, regional, district, constituency, sub-county, parish/ward and village/cell levels (Lotunani *et al*., 2014). It is recommended that both the health centres staffs and communities sit together when developing annual health work plans (Martineau *et al*., 2018). However, it is practically observed in most parts of Uganda that communities and other key stakeholders are not involved in making annual work plan for health centres (Lotunani *et al*., 2014; Muwanguzi *et al.,* 2020).

Annual health work plans can negatively or positively be associated with performance for the different health indicators (Martineau *et al*., 2018). The main aim of annual planning is to design well focused action plans that will be executed by the local government and related ministries (Lotunani *et al*., 2014). Health Centre Annual Work Plans are derived based on their allocation as routine and capital development budgets. Different health centres have been given opportunities to develop work plans since they experience different needs and priorities which has automatically exacerbated desirable support from funding bodies (Lotunani *et al*., 2014; O’Meara et al., 2011).

The annual health work plan is so much well-thought-out to have local government service delivery results including access to services by population, management of investment projects as per the guidelines through better leadership and commitment, resource mobilization and recruitment of staff, amongst others. Like other countries, discussions on services to be included in the work plan range from health education, to environmental determinants of health, to water and sanitation, to mental health and health-seeking behavior, amongst other (O’Meara et al., 2011).

Improved performance may be determined by the performance reports of health centres extracted from the annual work plans prepared and submitted to the district health office and ministry of health by March 31^st^ of every financial year. Importantly, management, monitoring and supervision of the work plans are necessary to account for district executions by the health centres’ budgets for routine services and investment projects (Local Government Management of Service Delivery Performance Assessment Manual, 2020). Lower community health centres smoothly implement their annual work plans when their health workers are highly motivated to perform their tasks and are given appropriate rewards (Lotunani *et al*., 2014).

The current assessment report by the prime minister’s office shows that annual health work plans in local governments have faced obstacles to being implemented every year (Local Government Management of Service Delivery Performance Assessment Report, 2020). There is therefore poorly made Annual Work Plans by the health facilities in Uganda (Ntirandekura et al, 2022). This is leading to poor health service delivery and causing slow progress to the achievement of the sustainable development goals. According to Lotunani *et al*. (2014), one of the obstacles to implementing the work plan is that leaders in the lower community health centres are not motivated and satisfied and this affects their health centre performance in the annual work planning process. Similarly, government grants sent to local governments indicate an inconsistency between lower health facilities proposed work plans and the total program delivery priorities funded by the district budget desk. Several lower health centres proposed work plans consistently continue to differ with the priority areas and budget allocations funded in the final consolidated districts’ budgets. Health centre leaders’ competence in health work planning is not being observed to give added value, particularly in supporting individual health centre and district performance (Lotunani *et al*., 2014).

This study determined the competences of lower community health centre leaders in annual health work plans and its influence on district performance improvement in the Busoga sub-region. It was proved important for top managers to ensure health centres meet staffing standards, actively participate in district planning cycles, and utilize a diverse set of implementation manuals for work planning to improve district performance.

### Research questions

1. What is the districts’ performance in annual health work planning in the Busoga sub-region?
2. What are the competences in annual health work planning of lower health facilities leaders that influence the performance of the districts?

### Objectives of the study

#### Broad objective

The broad objective of the study was to explore the competences of leaders of lower community health centres in annual health work planning that influence the districts’ performance in the Busoga sub-region for the last five years.

#### Specific objectives

1. To establish the districts’ performance in annual work planning in the Busoga sub-region.
2. To establish the influence of competence of the lower community health centre leaders in annual health work planning on the district performance in the Busoga sub-region.

## Methods

### Study design

This was a retrospective case control study conducted between July 2017 and June 2022. The reason for the use of a retrospective case control study design was to prove empirically and explain the effect of the competence in annual health work planning on the performance of districts in the Busoga sub-region. The case was districts with poor performance while the control was districts with good performance according to the ministry of health district performance league table and the local government performance assessment reports.

### Study Setting

The study was conducted in the Busoga sub-regions of eleven (11) districts and one (1) city. These include Mayuge, Namayingo, and Bugiri, Bugweri, Namutumba, Kaliro, Luuka, Kamuli, Buyende, Iganga, Jinja district local government and Jinja city.

### Variables and their measurements

The dependent variable is district performance and the independent variables are the competencies in annual health work planning for the different districts in the Busoga sub-region.

#### a) District performance in annual health work planning

Performance is normally looked at in terms of outcomes. According to Muwanguzi et al. (2020), performance can be looked at in terms of health seeking behaviour. Similarly, O’Meara et al. (2011) in Kenya agreed that performance indicators range from health education, to environmental determinants of health, to water and sanitation, to mental health and health-seeking behavior, amongst others. In Klassen et al. (2010) study, stated that employee’s performance is measured against the performance standards set by the organization. In Uganda, ministry of health has set performance standards in line with maternal child health, health education and promotion, environmental determinants of health, water and sanitation, mental health and health-seeking behavior, amongst others for the lower health centre leaders.

There are a number of measures that can be taken into consideration when measuring performance for example using of productivity, efficiency, effectiveness, quality and profitability measures (Adi, 2012). According to this study, performance was measured as a good one when three quarters (75%) of the priority areas identified in the facility work plan, are adopted and included in the final work plan of the district, there is an attached budget to every item and the level of funding in the work plan is realized. This is not far from the annual health sector performance reports for the selected five years where the national average score ranged from approximately 65 to 74 percent (approx. ^3^/_4_ of the performance mark). Any district which scored the national average mark and above was taken to be good performance.

#### b) Competence in annual health work planning

According to the study, competence is defined as the ability of the lower health centre leaders to successfully and effectively come up with an annual health work plan. The study established the influence of competence of the lower health centres’ leadership in annual health work planning on total program delivery in Busoga sub-region. According to O’Meara et al. (2011) competence requires knowledge and skills of the lower health centre leaders, aavailability of planning team and everyone’s involvement at the health centres, having timely planning meetings, and following the planning guidelines amongst others to clearly stipulate the competences of the leaders.

Qualified and motivated health workers are essential for adequate health service provision, however, it is still a challenge rural areas. Yet, according to the World Health Organisation (2006) performance especially in rural settings is considered to be a combination of staff being available all the time and staff being competent, productive and responsive. There is a statistically significant evidence of health work plans being useful in the implementation of activities generated by competent health workers in communities (Baker, 2004).

### Selection criteria of districts

The study considered the following;

1. District to have existed in the period of study.
2. Districts with performance below the national average (NA) for at least three years in the period 2017/18 to 2021/22 was considered for the case districts.
3. Districts with performance above the national average (NA) for at least four years in the period 2017/18 to 2021/22 was considered for the control districts.

Therefore, from the table, Namutumba, Namayingo, Mayuge and Luuka districts were picked for the case districts and Jinja, Iganga, Kamuli and Kaliro for the control districts.

### Sampling techniques

Purposive sampling was used to select the districts. Purposive sampling strategies are designed to enhance the understanding of selected individuals or groups’ experience(s) or for developing theories or concepts. Researchers seek to accomplish this goal by selecting “information-rich” cases, that is, individuals, groups, organizations, or behaviour (Devers & Frankel, 2000 cited in Nyaga, 2017).

Stratified random sampling technique was used to select lower public health centre IVs, IIIs and IIs for the study in Busoga sub region. Multistage stratified sampling was considered to determine the number of health centre IVs, IIIs and IIs selected for the public facilities in the case and control districts where data was collected.

### Study sample size determination

According to the District Health Information System 2 (DHIS2), the number of HCIV, III and II for each district and city in Busoga region are as reflected in table 2. Therefore, the total number of health centres from which the sampled health centres were picked is 330.

**Table 1:** Extract of District league table performance for the period 2017/18 to 2021/22.

| District | FY 2017/18<br>NA (69.2) | FY 2018/19<br>NA (73.1) | FY 2019/20<br>NA (68.1) | FY 2020/21<br>NA (64.5) | FY 2021/22<br>NA (68) |
| --- | --- | --- | --- | --- | --- |
| Bugiri | 73 | 70.7 | 72.09 | 64.7 | 60.9 |
| Bugweri | - | 64.5 | 72.74 | 53.3 | 58.8 |
| Buyende | 65.8 | 78.7 | 74.04 | 60.6 | 70.1 |
| Iganga | 72.1 | 74.9 | 73.53 | 66.8 | 67.2 |
| Jinja city | - | - | - | - | - |
| Jinja | 79.5 | 85 | 80.35 | 66.7 | 64.6 |
| Kaliro | 71 | 66.3 | 71.36 | 70.1 | 68 |
| Kamuli | 70.9 | 74.6 | 71.96 | 66.1 | 65.3 |
| Luuka | 56.3 | 68.7 | 65.94 | 63.1 | 65.9 |
| Mayuge | 63.7 | 66.3 | 70.58 | 55.7 | 58.7 |
| Namayingo | 66.2 | 70.7 | 70.21 | 60.9 | 66.7 |
| Namutumba | 73.7 | 72.4 | 59.29 | 60.2 | 61.5 |

Source: Annual health sector performance reports; 2017/18, 2018/19, 2019/20, 2020/21 and 2021/2022

**Table 2:** Number of health centre IV, III & II for districts in Busoga sub region.

| Districts | Level of Health Centre |  |  |  |
| --- | --- | --- | --- | --- |
|  | IV | III | II | Overall total |
| Bugiri | 1 | 10 | 26 | 37 |
| Bugweri | 1 | 3 | 12 | 16 |
| Buyende | 1 | 4 | 12 | 17 |
| Iganga | 1 | 9 | 18 | 28 |
| Jinja city | 4 | 8 | 13 | 27 |
| Jinja | 1 | 7 | 24 | 32 |
| Kaliro | 1 | 6 | 7 | 14 |
| Kamuli | 2 | 12 | 26 | 40 |
| Luuka | 1 | 6 | 22 | 29 |
| Mayuge | 3 | 9 | 28 | 40 |
| Namayingo | 1 | 5 | 21 | 27 |
| Namutumba | 1 | 4 | 18 | 23 |
| Total | 18 | 83 | 227 | 330 |

Source: DHIS2 21 March, 2023

Considering Muwanguzi, et al. (2020) study on the effectiveness of training of health unit management committee (HUMC) members, quoted Schelssman’s (1982) formula to determine the sample size of health centres.

Where n =the number of the required sample size of the case. Z_α_is thestandard normal value corresponding to the required level of significance for 0.05 (1.96),Z_ß_ is the standard normal value corresponding to required power of study for 80% (0.84), P_0_ isthe probability of success among the control districts and P_1_ is the probability of success among the case districts. However, _C_ = 1, which is the number of controls per case, and Q = 1 – P_1_. According to May (2018) study on the implementation of grip strength measurement in medicine for older people wards as part of routine admission assessment; identifying facilitators and barriers using a theory-led intervention in Newcastle, UK where P_0_ was 46.3% and P_1_ 79%.

Number (n) of health centres for the case distircts was 12 and 12 health centres for the control districts. The 12 health centres in each of the district categories (good or poorly performing) was randomly selected for data collection.

See table 3 showing the actual number of health centres at the different levels selected per district.

**Table 3:** Number of health centre IV, III & II selected for the study in the four districts.

| Performance of districts | Level of Health Centre |  |  |  |  |
| --- | --- | --- | --- | --- | --- |
|  |  | IV | III | II | Total |
| Poor performance | Overall number of HCs | 5 | 34 | 85 | 124 |
|  | Sampled number of HCs | 1 | 3 | 8 | 12 |
| Good performance | Overall number of HCs | 6 | 24 | 89 | 119 |
|  | Sampled number of HCs | 1 | 3 | 8 | 12 |

### Data collection techniques

#### a) Documentary Review

Under the secondary collection of data, documented records provide the necessary background and a lot of necessary information for a more organized work (Kothar, 2004). Where possible, depending on the cooperation of district managers and health centre leaders, the researcher reviewed the annual health work plan at the health centres to understand the work plan processes, its submission, its contents and adoption by the district budget desk, performance management records and others. The data collection process took place between 17^th^ July, 2024 and 23^rd^ August, 2024. The review concentrated much on maternal child health indicators, health education and promotion, water and sanitation and other environmental health related indicators, mental health and health-seeking behavior, infrastructure, amongst others.

#### b) Administering of questionnaire

Leaders were asked questions to gain information on their competences as supported by Amin (2005). This was a questionnaire with different sections of questions answered by leaders at the health centres. The appointments with lower community health centre leaders were made to avoid missing the leader at the health centres visited.

### Research instruments

Clifton & Handy (2001) stated that “choosing among the different data collecting tools involves considering their appropriateness and relative strengths and weaknesses.” In this study, combinations of tools were used, that is, records review checklist and questionnaire. These tools were designed using the key study themes/objectives. The secondary data collection tools included work plans for the last five years at health centres and the annual assessment service delivery performance reports at the districts.

#### a) Questionnaire

A questionnaire is a set of written questions stemming from the objectives of the study and literature review which is developed by the researcher and administered to a selected group of respondents (Clifton & Handy, 2001). It is more preferable especially when dealing with respondents who do not offer time for interview and are very free and open when it comes to putting their views on paper. Questionnaires were administered to the consenting lower community health centre leaders. Questionnaires were filled by the research assistants for respondents who did not want to write for themselves.

#### b) Document / Records Review Checklist

A checklist will be a tool designed to capture information extracted from the annual health work plans and the annual assessment service delivery performance reports at the districts for the previous five years.

### Data quality control

#### a) Validity of Instruments

Validity involves getting the most accurate data. It is defined as the degree to which a test or measuring instrument measures what it purports to measure, or how well a test or an instrument fulfils its function (Anastasi & Urbina, 1997). Content Validity Index will be used to get the validity of the research instruments.

Content Validity Index (CVI) was performed based on items derived from instruments and volunteer evaluators. Each evaluator will rate the questions on a two-point rating scale *of **Relevant (R)*** and ***Irrelevant (IR)***. Thereafter, the computation of CVI will be done by summing up the judges’ ratings on either side of the scale and dividing the two to obtain the average.

#### b) Reliability of instruments

The researcher will pre-test the instrument to ascertain the level of consistency, weakness, and unclear questions to 10 respondents and adjustments done to enhance its reliability. The researcher will use Cronbach’s alpha coefficient on analysis to identify the only variables with an alpha coefficient of more than 0.70 which is acceptable for social research (Amin, 2005).

### Data analysis

Data analysis was computed using STATA *version* 15 for the quantitative data. Tests of independence was used to determine the statistical significance of different variables. The *p*-value set at 0.05 will be used to determine the statistical significance of the associations between independent and dependent variables at 95 percent confidence intervals (CI).

Logistic regression model was used to ascertain the statistical relationship between independent and dependent variables. It will derive regression odds ratios and p-values which was used to determine which variables impacted on the topic of study. Furthermore, multiple logistic regression of independent variables was considered to confirm whether they were the predictors of performance improvement.

### Ethical approval and consent to participants

Ethical approval to conduct the study was provided by the Institutional Review Boards (IRB) of Uganda Martyrs University - Nsambya hospital (SFHN – 2024 – 134). Written consent was obtained from all the study participants and authorities in the study area. Privacy and confidentiality of the information was assured to the respondents. The participants consented to participate in the study and were assured of their right to withdraw from the engagement at their will. This is a key aspect of the consenting process for the study participants.

## Results

The results from the study present the performance assessment of health facilities across five financial years in Busoga sub-region. The findings show the performance scores by facility level and the overall performance of the case and control health centres. A total 24 health centres (12 case health centres and 12 control) constituting of 2 HCIVs, 6 HCIIIs, and 16 HCIIs, were studied across five different financial years giving 120 possible observations.

### Districts’ performance in annual health work planning in Busoga sub-region

Performance of health facilities was assessed against the facility having planned for maternal and child health activities; health promotion, disease prevention, sanitation and hygiene activities; administrative activities; infrastructure activities; and routine mobilization activities, performed routine performance and consequent improvement activities (quarterly performance reviews and implementation report). In addition, number of people involved in the annual planning meetings (4 and more times is better performance), number of meetings attended by persons involved in annual health work planning (4 and more times is better performance) and tools used for planning (3 and more tools used is better performance).

Each correct activity was scored 1, otherwise 0. A health centre that correctly did 75% of the activities was graded as good performance otherwise poor performance. Overall, the performance for each health centre level across the five financial years showed that poor performance in all the participating health centres, though control health centres performed better than the case health centres. The overall performance of health centres in the control was 47.2% and in the case 26.4%, with an average of 49.1% among the control health centre IV compared to 18.2% among case health canter IV. Also, 48.5% performance was among control health centre IIIs compared to 35.8% among case health centre IIIs. Similarly, HCII of the control had an average performance of 43.9% compared to 25.2% of the case HCIIs. The detail of each financial year variations among the different level of the health facilities was present in table 4 and figure 1.

**Figure 1:**
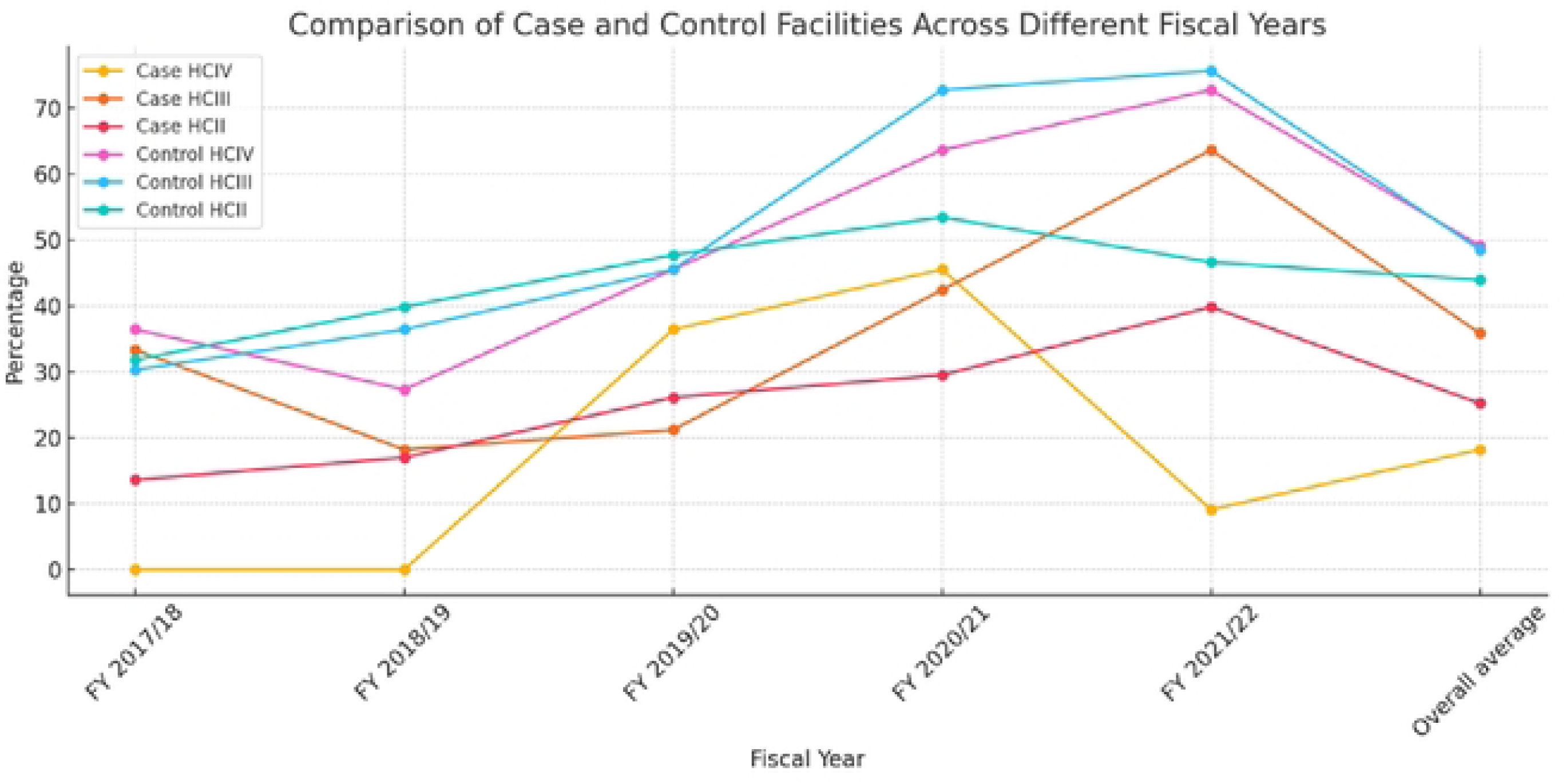
comparison of the case and control health centres at different levels across different financial year.

**Table 4:** Performance of the health facilities in Busoga sub-region across 5 fiscal years period.

| Facility level | FY 2017/18 | FY 2018/19 | FY 2019/20 | FY 2020/21 | FY 2021/22 | Overall average |
| --- | --- | --- | --- | --- | --- | --- |
| Case facilities |  |  |  |  |  | **26.4** |
| HCIV | 0 | 0 | 36.4 | 45.5 | 9.1 | 18.2 |
| HCIII | 33.3 | 18.2 | 21.2 | 42.4 | 63.6 | 35.8 |
| HCII | 13.6 | 17.0 | 26.1 | 29.5 | 39.8 | 25.2 |
| Control facilities |  |  |  |  |  | **47.2** |
| HCIV | 36.4 | 27.3 | 45.5 | 63.6 | 72.7 | 49.1 |
| HCIII | 30.3 | 36.4 | 45.5 | 72.7 | 75.6 | 48.5 |
| HCII | 31.8 | 39.8 | 47.7 | 53.4 | 46.6 | 43.9 |

But also the study discovered that at the different levels of health centres (HCIVs, HCIIIs and HCIIs), control facilities performed better than the case facilities (see figure 2). However, their performance was not the desired one (75 percent performance was not realised). Equally, the overall performance of health centres in the control and case was not the desired one as reflected in figure 3.

**Figure 2:**
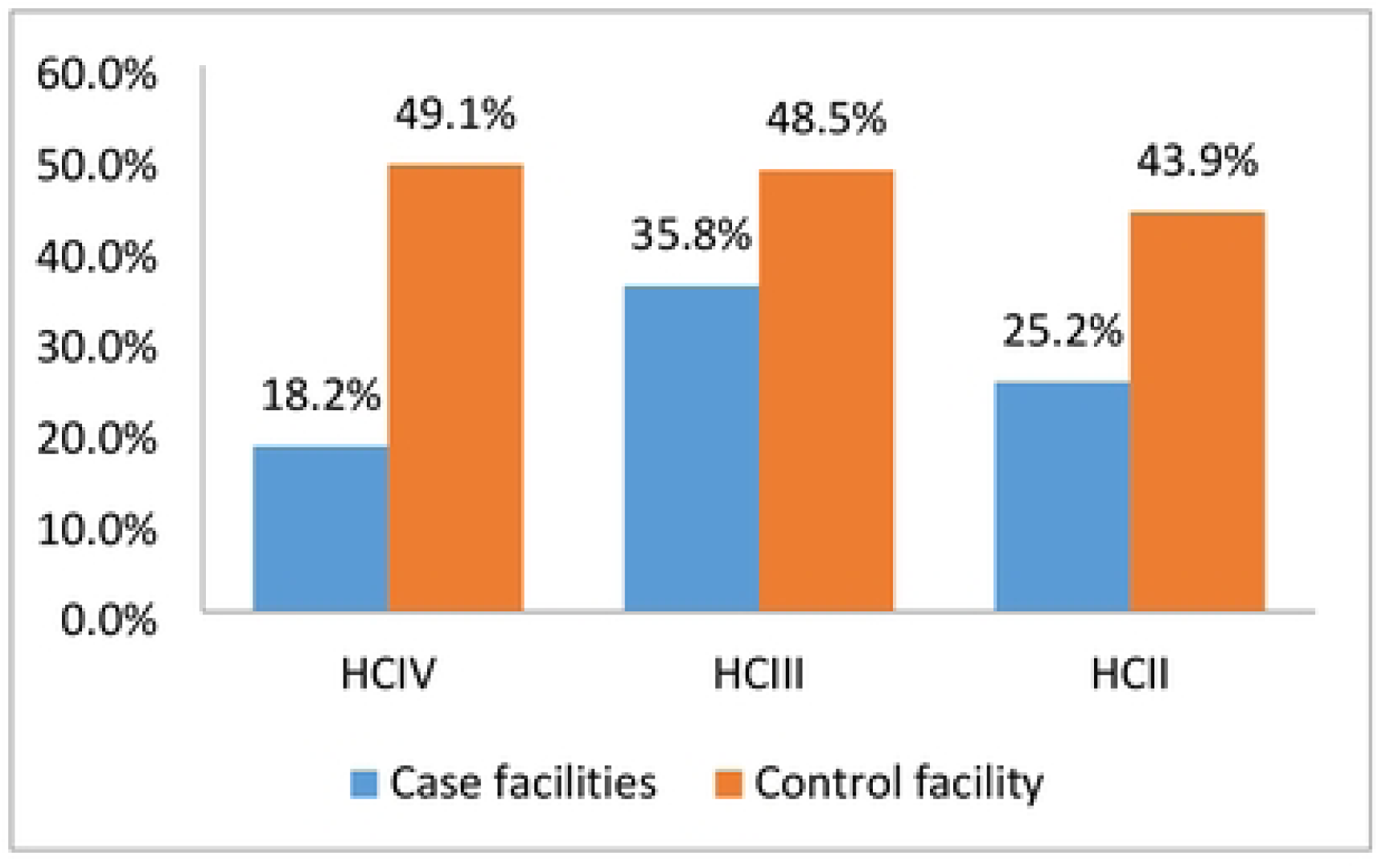
Showing the comparison between case and control facilities

**Figure 3:**
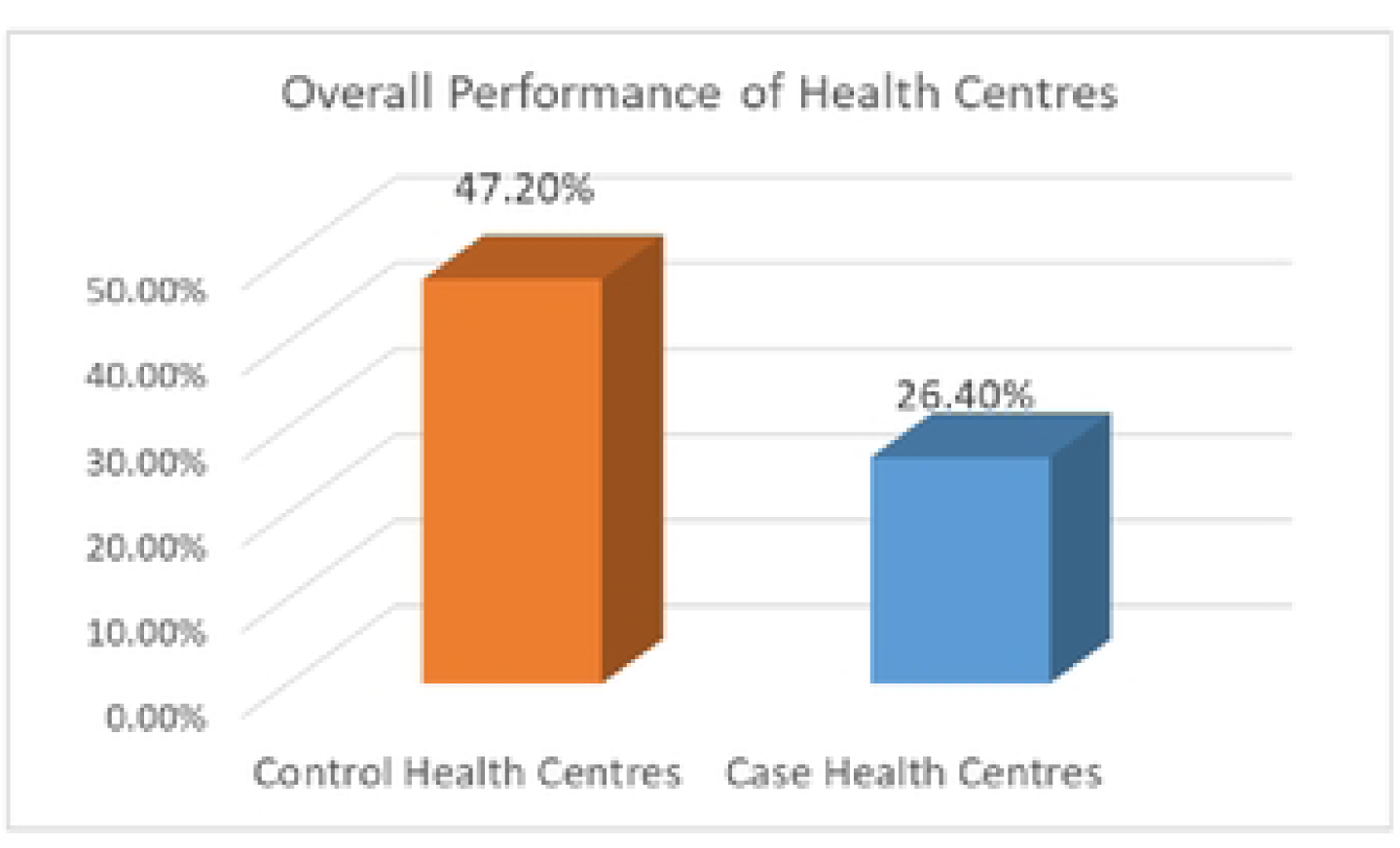
Showing the overall performance in case and control health centre*s*

The performance trend of the health centres in the control and case over the 5 years is presented in figure 4. Generally, the control facilities performed better than the control facilities in the subsequent five years. Important to note is the gradual improvement in performance for the two groups which was observed especially in the last four years.

**Figure 4:**
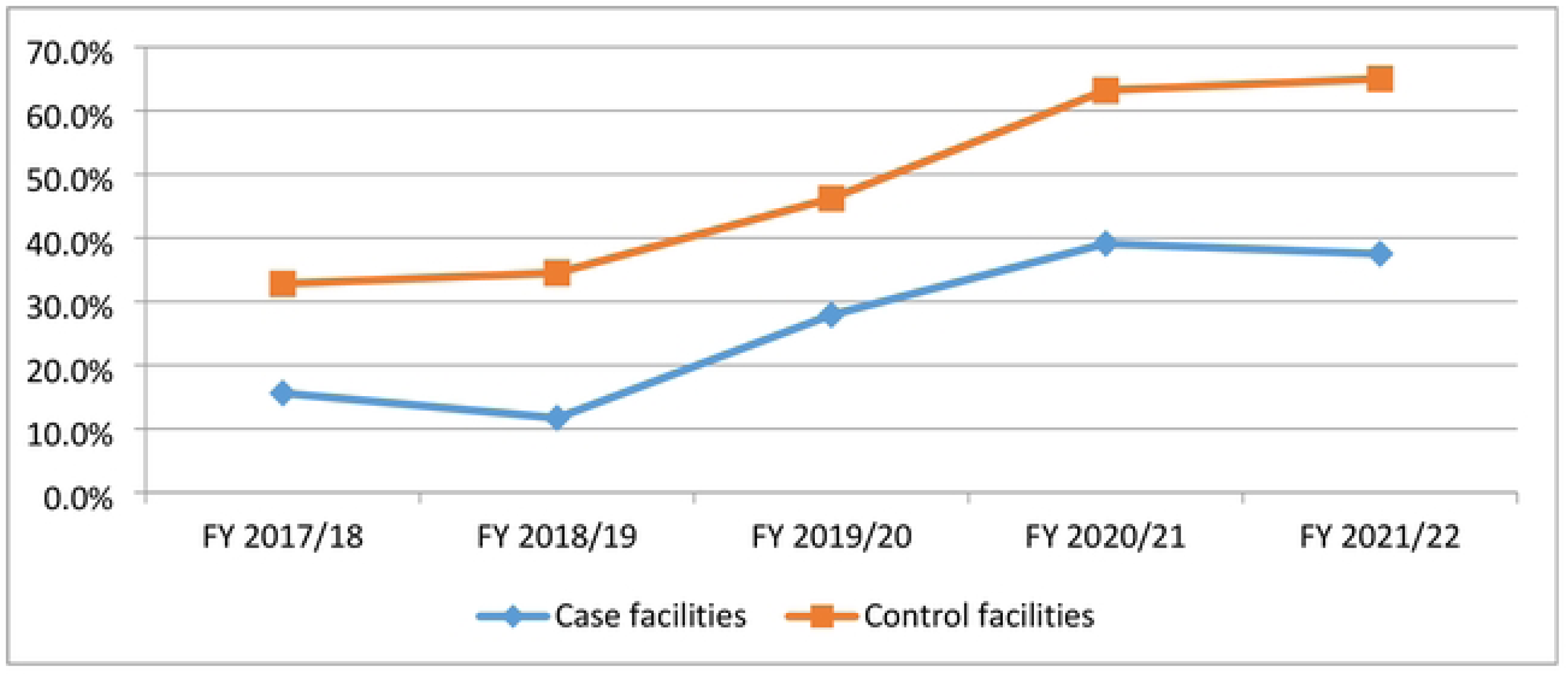
showing the trend of performance of health centres in case and control.

### Competences of leaders of lower community health centres in annual health work planning

Table 2 indicates that all the health centres (both case and control) 100% had HUMC, 90% of the control facilities had the HUMC fully constituted while 50% of the case facilities had the HUMC fully constituted. About three quarter of all health centres had HUMC members meeting the academic requirements. Only 13.3% and 10% of the control and case health centres had the staffing level meeting the standards for the health centres respectively. The table provides more detail about the competence activities.

For the 5 years, 80 percent of the health centres in the control and 83.3 percent in the case had not attended 4 or more planning cycle meetings organised by the district in the five years. Equally, most health centres did not organise annual health work planning meetings at the facility level, and less than 4 people were involved in the planning process of the work plan (43.3% for the controls and 70% for the cases). There are many tools which are used to guide the planning process, but only 13.3% health centres in both the control and case were able to use at least three or more tools for better planning as shown in table 5.

**Table 5:** Showing the competence of the health facilities.

| Variables/Activities | Control | Case |
| --- | --- | --- |
|  | Freq. (%) | Freq. (%) |
| There a HUMC |  |  |
| Yes | 60 (100) | 60 (100) |
| No | 0 (0.0) | 0 (0.0) |
| HUMC fully constituted |  |  |
| Yes | 56 (90) | 30 (50.0) |
| No | 4 (10) | 30 (50.0) |
| HUMC members meeting the academic requirements |  |  |
| Yes | 45 (75) | 46 (76.7) |
| No | 15 (25) | 14 (23.3) |
| Staffing level meets the standards for the facility |  |  |
| Yes | 8 (13.3) | 6 (10) |
| No | 52 (87.7) | 54 (90) |
|  | Freq. (%) | Freq. (%) |
| Ever attended planning cycle meetings organized by the district health department |  |  |
| Yes | 12 (20) | 10 (16.7) |
| No | 48 (80) | 50 (83.3) |
| Number of planning cycle meetings organized by the district health department you attended. |  |  |
| Attended at least 4 times for the 5 years | 12 (20) | 10 (16.7) |
| Attended less than 4 times for the 5 years | 48 (80) | 50 (83.3) |
| Organized annual planning meetings at the facility |  |  |
| Yes | 34 (56.7) | 18 (30) |
| No | 26 (43.3) | 42 (70) |
| Persons involved in the annual planning process |  |  |
| 4 and more persons involved | 34(56.7) | 18 (30) |
| < 4 persons involved | 26(43.3) | 42 (70) |
| Tool used for developing annual health work plan: |  |  |
| 3 and more tools mentioned | 8 (13.3) | 8 (13.3) |
| < 3 tool mentioned | 52 (86.7) | 52 (86.7) |

### Influence of competences of the lower health centre leaders on the districts performance in the Busoga sub-region

When the different competence activities / factors were assessed against overall performance level, the following competence variables influenced the district performance and were statistically significant (*p-value* less than 0.05); having a staffing level that meets the standard of public health centres, having the health centre attending / represented in a planning cycle meetings organized by the district health departments and lastly using at least four and more implementation manuals to develop meaningful annual health work plans. The details are as shown table 6 below.

**Table 6:** Influence of competence of the lower health centre leaders in annual health work planning on the performance in the Busoga sub-region 2017/18-2021/22.

| Variables | $\chi^2$ | p-value |
| --- | --- | --- |
| There a HUMC |  |  |
| Yes |  |  |
| HUMC fully constituted | 0.741 | 0.389 |
| Yes |  |  |
| No |  |  |
| HUMC members meeting the academic requirements | 2.222 | 0.136 |
| Yes |  |  |
| No |  |  |
| Staffing level meets the standards for the facility | 7.756 | **0.005\*** |
| Yes |  |  |
| No |  |  |
| HODs involved in the annual planning process | 0.190 | 0.663 |
| Yes |  |  |
| No |  |  |
| HUMCs involved in the annual planning process | 2.963 | 0.085 |
| Yes |  |  |
| No |  |  |
| Ever attended planning cycle meetings organized by the district health department | 16.713 | **0.001\*** |
| Yes |  |  |
| No |  |  |
| Organized annual planning meetings at the facility | 1.931 | 0.165 |
| Yes |  |  |
| No |  |  |
| At least 4 implementation manual used use to plan | 43.333 | **0.001\*** |
| Yes |  |  |
| No |  |  |

## Discussion

The findings of this retrospective study indicate that specific competences of leaders in lower-level community health centres significantly influence the performance improvement of districts in the Busoga sub-region. Notably, three key competences emerged as significant contributors to district performance:

1. Staffing levels to meet public health centre standards was significant for this study. Unfortunately, in this study, about 13 percent of health centres had 100 percent staffing. Health centres with adequate staffing that meet public facility standards demonstrated a higher capacity to implement effective health services. Lotunani *et al*. (2014) says focussed planning and staff recruitment inclusive enables local governments perform. Adequate staffing allows for the distribution of workload during the planning phase, proper delegation of duties, and efficient service delivery, which collectively improve health outcomes. The positive relationship between appropriate staffing levels and district health performance emphasizes the importance of human resource management in achieving public health goals. Staffing level at 100 percent is a requirement for every health centre according to the Local Government Management of Service Delivery Performance Assessment Manual (2020).
2. Participation in planning cycle meetings is very necessary to health centre leaders for proper planning. Ministry of Finance, Planning and Economic Development (MoFPED) is always organising regional planning meetings for each financial year. Regional meetings are attended by departmental heads and political representatives purposely to receive guidelines for use during planning and budgeting period (MoFPED, 2022). Health centres that actively participated or were represented in planning cycle meetings organized by district health departments showed a significant improvement in performance. However, about 20 percent of lower community health centre leaders attended the planning meeting. This clearly reflects the failure of departmental heads to implement the planning for health centres based on the existing guidelines. Perhaps it is attributed to lack of trained staff and financial constraints as stated by O’Meara et al. (2011). Community representatives equally did not attend these meetings. In Kenya, community perspective was brought by the facility committee, in which each village is intended to be represented by a committee member during the planning period in addition to the health centre in-charges (O’Meara et al., 2011). Involvement in these meetings ensures that local health centres are aligned with district-wide health priorities, resource allocations, and implementation strategies. Like in Kenya, the committees discussed their inputs in the work plan and the barriers to health at each life stage based on the guidelines which were issued by the MoH (O’Meara et al., 2011). This alignment is crucial for the effective coordination of healthcare delivery, enabling lower-level centres to contribute meaningfully to district-wide health improvement initiatives.
3. Use of Implementation Manuals for Developing Annual Health Work Plans was significant noted for this study. The use of at least four or more implementation manuals in the development of annual health work plans was found to significantly influence district performance. The study concur with O’Meara et al. (2011) and World Health Organisation (2020) that guidelines issued by the MoH are important when developing annual health work plans. Manuals provide structured guidelines that ensure health centres adhere to national and district-level standards when formulating their health work plans. These manuals include the PHC guidelines, RBF guidelines, procurement guideline amongst others which often cover areas such as service delivery protocols, resource management, and community engagement that are critical for developing comprehensive and actionable work plans. Committees in Kenya discussed pregnancy and new-born, early childhood, late childhood, adolescence, and adulthood at facility and community barazas (public meetings) as per the guidelines (O’Meara et al., 2011), not far from this study. Centres that utilized a variety of manuals were able to implement more meaningful and evidence-based health interventions, leading to better health outcomes at the district level.

The combination of these factors suggests that the competence of lower-level health centre leadership, particularly in the areas of staffing, participation in district planning processes, and adherence to standardized guidelines, plays a pivotal role in improving district health performance. It highlights the need for continuous capacity building for health centre leaders to ensure they have the skills and resources necessary to contribute effectively to district health objectives.

## Conclusion

This study underscores the critical role of lower-level community health centre leaders in driving district health performance improvement in the Busoga sub-region. Health centres that meet staffing standards, actively participate in district planning cycles, and utilize a diverse set of implementation manuals for work planning are better positioned to contribute to district-wide health improvements. These findings suggest that investing in leadership competences at the community health centre level is essential for achieving district health performance goals. Additionally, the study provides empirical evidence that strengthening these key competences can lead to statistically significant improvements in health outcomes at the district level.

### Recommendations

1. Strengthening Staffing Levels: It is recommended that the Ministry of Health and district health departments prioritize maintaining adequate staffing levels at lower-level community health centres. This may include recruitment drives, retention strategies, and periodic assessments to ensure staffing standards are met.
2. Enhancing Participation in District Health Planning: Health centre leaders should be encouraged and supported to participate in district-level planning cycle meetings. This could be facilitated by providing transport allowances, capacity-building workshops, and ensuring that planning meetings are scheduled at times that do not conflict with other duties.
3. Promoting the Use of Implementation Manuals: District health departments should ensure that lower-level health centres have access to updated and comprehensive implementation manuals. Regular training on the use of these manuals should be provided to ensure that health centre leaders can develop robust and effective annual health work plans.
4. Capacity Building for Health Centre Leaders: Continuous professional development opportunities should be made available to health centre leaders, focusing on leadership, health planning, and resource management. Such training will empower leaders to make informed decisions that positively influence district health performance.

## Data Availability

data will be available on request

## Abbreviations

AWP: Annual Work Plans

LLGs: Lower Local Governments

NA: National Average

FY: Financial Year

DHIS2: District Health Information System 2

HC: Health Centre

HUMC: Health Unit Management

CVI: Committee Content Validity Index

CI: Confidence Interval

IRB: Institutional Review Boards

MoFPED: Ministry of Finance Planning and Economic Development.

## Acknowledgements

The authors extend their sincere appreciation to Paul Kitakule, Mohammed Mukalu, District Health Officers, Biostatisticians and In-charges of health centres for their support in data collection. We are also grateful to the study participants, lower community leadership and the centre staff in the study region for their contributions to this study.

## Competing interests

The authors declare that they have no competing interest.

## Authors’ contributions

KMM conceived the study idea. NR, KMM, JFM, MW, PW, and OGO designed the study and wrote the protocol. KMM, and DGM designed the data collection tools. KMM, IW, DGM participated in data collection. KMM, DGM, MW, PK and BA undertook the analysis. KMM, DGM, PW, JFK and BA wrote the manuscript.

## Funding

There was no external funding for this study. All funding was contributed by the authors from their respective work places.

## Notes

### Competing Interest Statement

The authors have declared no competing interest.

### Funding Statement

The author(s) received no specific funding for this work.

### Author Declarations

Ethical approval to conduct the study was provided by the Institutional Review Boards (IRB) of Uganda Martyrs University – Nsambya hospital (SFHN — 2024 – 134).

